# Fine-scale heterogeneity in *Schistosoma mansoni* force of infection measured through antibody response

**DOI:** 10.1101/2020.04.10.20061101

**Authors:** Benjamin F. Arnold, Henry Kanyi, Sammy M. Njenga, Fredrick O. Rawago, Jeffrey W. Priest, W. Evan Secor, Patrick J. Lammie, Kimberly Y. Won, Maurice R. Odiere

## Abstract

Identifying populations with active transmission and monitoring changes in transmission is centrally important in guiding schistosomiasis control programs. Traditionally, human *Schistosoma mansoni* infections have been detected in stool using microscopy, which is logistically difficult at program scale and has low sensitivity when people have low infection burdens. We compared serological measures of transmission based on antibody response to schistosomiasis soluble egg antigen (SEA) with stool-based measures of infection among 3,663 preschool-age children in an area endemic for *S. mansoni* in western Kenya. Serological measures of transmission closely aligned with stool-based measures of infection, and serological measures provided better resolution for between-community differences at lower levels of infection. Serology enabled fine-scale measures of heterogeneity in force of infection both geographically and by age. Our results show that serologic surveillance platforms represent an important new opportunity to guide and monitor schistosomiasis control programs.

## Introduction

Schistosomiasis is one of the most common parasitic diseases in the world, with an estimated 142 million people infected globally, over 90% of whom live in Sub-Saharan Africa.^1,2^ The disease is caused by infection with blood flukes of the genus *Schistosoma* spp. Adult *Schistosoma mansoni* worms live in the mesenteric veins adjacent to the human intestinal tract. *S. mansoni* is highly endemic in the Lake Victoria region, where dermal contact with the cercariae stage of the parasite in surface waters is the route of primary transmission.^3^

The World Health Organization has targeted the global elimination of schistosomiasis as a public health goal by 2025, defined as a reduction to <1% prevalence of heavy intensity infections across sentinel sites.^2^ A cornerstone of the global control strategy has been delivery of the drug praziquantel to school-age children (5 – 15 years) through mass drug administration (MDA) campaigns in endemic regions.^4^ School-age children have been a focus of MDA efforts because *S. mansoni* infection intensity and egg shedding peaks at ages 10 – 15 years,^3^ and because schools provide a logistically convenient approach for locating children through large-scale programs. Measuring *S. mansoni* infection among children is crucially important to target program efforts to populations with ongoing transmission and to monitor control program progress. Monitoring transmission requires sensitive markers of infection that can be measured in large-scale surveys.

Seroepidemiologic studies that measure transmission intensity through antibodies in blood have made valuable contributions to the study of transmission dynamics and burden of infectious diseases such as malaria,^5,6^ arboviruses,^7–10^ and enteric pathogens.^11–13^ Serological testing is growing in use to monitor progress of many neglected tropical disease control programs such as trachoma,^14^ lymphatic filariasis,^15^ and onchocerciasis.^16^ The near-term potential for integrated, population-based serologic surveillance with multiplex assays creates an opportunity for more frequent, broader monitoring of diseases with well-validated serological assays and methods to translate antibody response into measures of transmission.^17,18^ To our knowledge, there has been no detailed assessment of seroepidemiologic methods used to estimate transmission based on *S. mansoni* antibody measurements, nor has there been a thorough comparison of serological measures of transmission against the current standard of stool-based measures of patent infection.

Our objective was to estimate *S. mansoni* transmission through antibody responses measured among 3,663 pre-school age children living in 30 villages in western Kenya near Lake Victoria, an area endemic for *S. mansoni*. We studied the relationship between serologic measures and stool-based measures of transmission intensity and used serology to create high resolution assessments of force of infection by distance from Lake Victoria and by age. Our results show that population-based, serological surveys create a new opportunity to monitor *S. mansoni* transmission and control program progress in endemic settings and further suggest that preschool age children represent an important sentinel population for serologic monitoring in higher transmission settings.

## Results

### Study population and setting

The analysis included 3,663 children ages 2 months to 5.5 years who were originally enrolled from 30 communities that participated in a cluster randomized controlled trial to measure the effect of community-wide versus school-based mass distribution of praziquantel to reduce *S. mansoni* infection in Mbita, western Kenya.^19^ Villages were selected within 5 km of Lake Victoria from among those with ≥ 25% *S. mansoni* prevalence ^20^ that had not received mass treatment with praziquantel. Preschool age children ≤5 years were assessed annually from 2012-2014 in repeated cross-sectional surveys that collected blood and stool specimens, and samples were tested for the presence of infection (in stool) and for antibodies to *S. mansoni* soluble egg antigen (SEA). The area is highly endemic for malaria and schistosomiasis infection.

*S. mansoni* infection prevalence decreased slightly over the study period (Table 1), but was not significantly different by treatment arms,^19^ so we combined data from all years (2012-2014) in the present analysis. The average number of children measured per community during the three-year study period was 122 (median: 117, inter-quartile range: 84, 163; range: 59, 196). *S. mansoni* infection and seroprevalence increased with age, consistent with endemic transmission (Table 1). Model-based geostatistical predictions of seroprevalence as a function of location plus a range of environmental covariates showed that infection was highest close to Lake Victoria, with some heterogeneity over the study area (Fig 1). Predicted prevalence of stool-based infection showed a similar pattern and were consistent with geospatial analyses of infection among school-age children in the region (Supplementary Fig 1).^21^

**Table 1.** Number of samples tested and *Schistosoma mansoni* prevalence by soluble egg antigen (SEA) and Kato-Katz, stratified by age and by year, Mbita, Kenya, 2012- 2014.

|  | SEA Seroprevalence |  |  | Kato-Katz Prevalence |  |  |
| --- | --- | --- | --- | --- | --- | --- |
|  | N<br>Samples | N<br>Positive | (%) | N<br>Samples | N<br>Positive | (%) |
| **Overall** | 3,663 | 1,749 | 48 | 3,426 | 869 | 25 |
| **Age, years completed** |  |  |  |  |  |  |
| <1 year | 51 | 12 | 24 | 47 | 5 | 11 |
| 1 year | 570 | 132 | 23 | 503 | 68 | 14 |
| 2 years | 780 | 295 | 38 | 714 | 150 | 21 |
| 3 years | 890 | 483 | 54 | 844 | 211 | 25 |
| 4 years | 1,214 | 727 | 60 | 1,166 | 384 | 33 |
| 5 years | 158 | 100 | 63 | 152 | 51 | 34 |
| **Year** |  |  |  |  |  |  |
| 2012 | 1,120 | 557 | 50 | 1,072 | 297 | 28 |
| 2013 | 1,187 | 595 | 50 | 1,172 | 308 | 26 |
| 2014 | 1,356 | 597 | 44 | 1,182 | 264 | 22 |

**Figure 1.**
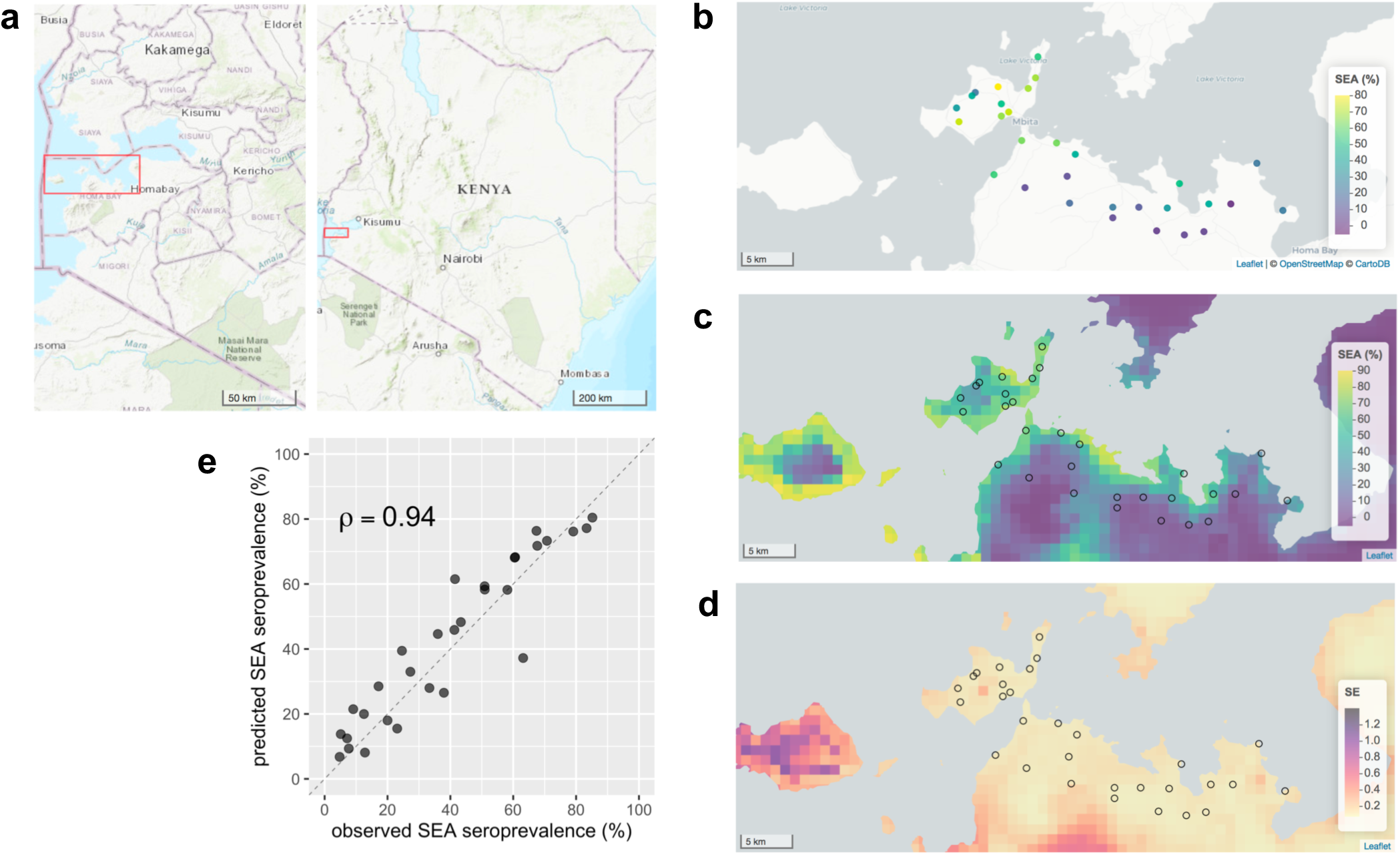
Spatial heterogeneity of antibody response to *Schisotosoma mansoni* SEA antigen in 30 study communities near Mbita, Kenya, 2012–2014. (**a**) Overview of the study location. Red rectangles mark the extent of the remaining map panels. (**b**) SEA seroprevalence in the 30 study communities measured from 3,663 preschool aged children. (**c**) Predicted seroprevalence at 1 km resolution from a geostatistical model. (**d**) Approximate standard errors of the predicted proportion SEA seropositive from the geostatistical model. (**e**) Geostatistical model predicted SEA seroprevalence versus observed for the 30 study communities in 2014. Spearman rank correlation (*ρ*) estimate between predicted and observed. The diagonal line is 1:1. Created with notebook: https://osf.io/eqp3w/.

### Comparison of serological and stool-based measures of transmission

Community-level seroprevalence was consistently higher than infection prevalence detected by stool exam, but measurements were highly correlated (Spearman’s *ρ* = 0.95) and the relationship was consistent across study years (Fig 2a). At the community level, we observed a wider range of seroprevalence at low levels of stool-based prevalence, suggesting that serology had higher sensitivity and better resolution compared with stool-based testing at lower levels of transmission.

**Figure 2.**
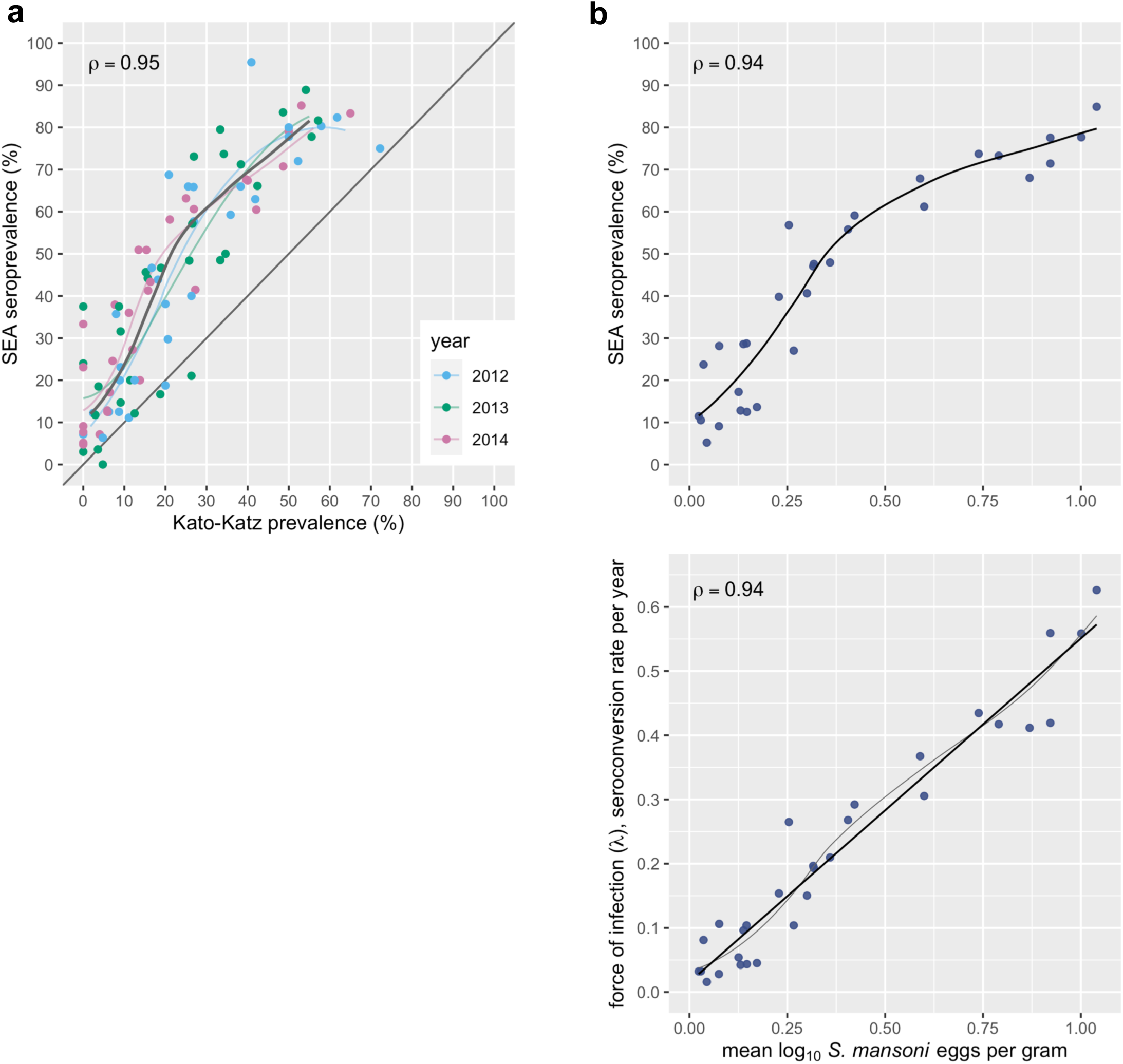
Community-level seroprevalence of *Schisotosoma mansoni* SEA response and infection measured by double-slide Kato-Katz stool microscopy in Mbita, Kenya, 2012–2014. (**a**) Relationship beween SEA seroprevalence and Kato-Katz prevalence in the 30 study communities over a three year period. Locally weighted regression fits are provided for each year, along with a single fit from measurements pooled across years (heavy line). Regression fits trimmed to 95% of the data to reduce edge effects. (**b**) Community level *S. mansoni* SEA seroprevalence and force of infection as a function of mean log_10_ *S. mansoni* eggs per gram of stool, a conventional measure of infection burden. Panels include locally weighted regression fits; the force of infection panel also includes a linear fit (heavy line). Each panel includes Spearman rank correlation estimates (*ρ*). Created with notebook: https://osf.io/mhdjx/.

Community-level seroprevalence and infection prevalence was relatively stable over the 3-year study period, with high correlation between years (Supplementary Fig 2) even in the context of ongoing MDA. Preschool age children were not directly targeted in the study’s MDA campaigns, but treatment with praziquantel was offered for all study children with positive stool microscopy.

At the community level, serological measures of transmission intensity aligned closely with stool-based measures of infection intensity. We measured a child’s *S. mansoni* infection intensity using eggs per gram of stool which, at the population level, has been used as a measure of parasite burden.^22^ We also estimated each community’s average force of infection with the seroconversion rate (details in Materials and Methods). There was a monotonic relationship between community mean infection intensity and seroprevalence, and a linear relationship between mean infection intensity and force of infection (Fig 2b).

Simulations that varied the sample size per community between 20 and 200 measurements showed that the correlation remained strong between summary measures of SEA or Kato-Katz prevalence and community level force of infection estimated in the full sample (Fig 3). In the present setting, we estimated serological surveys that measure as few as 20-40 children per community could reliably rank communities according to their underlying transmission intensity (*ρ*>0.9), similar to estimates for dengue seroprevalence in Bangladesh.^8^

**Figure 3.**
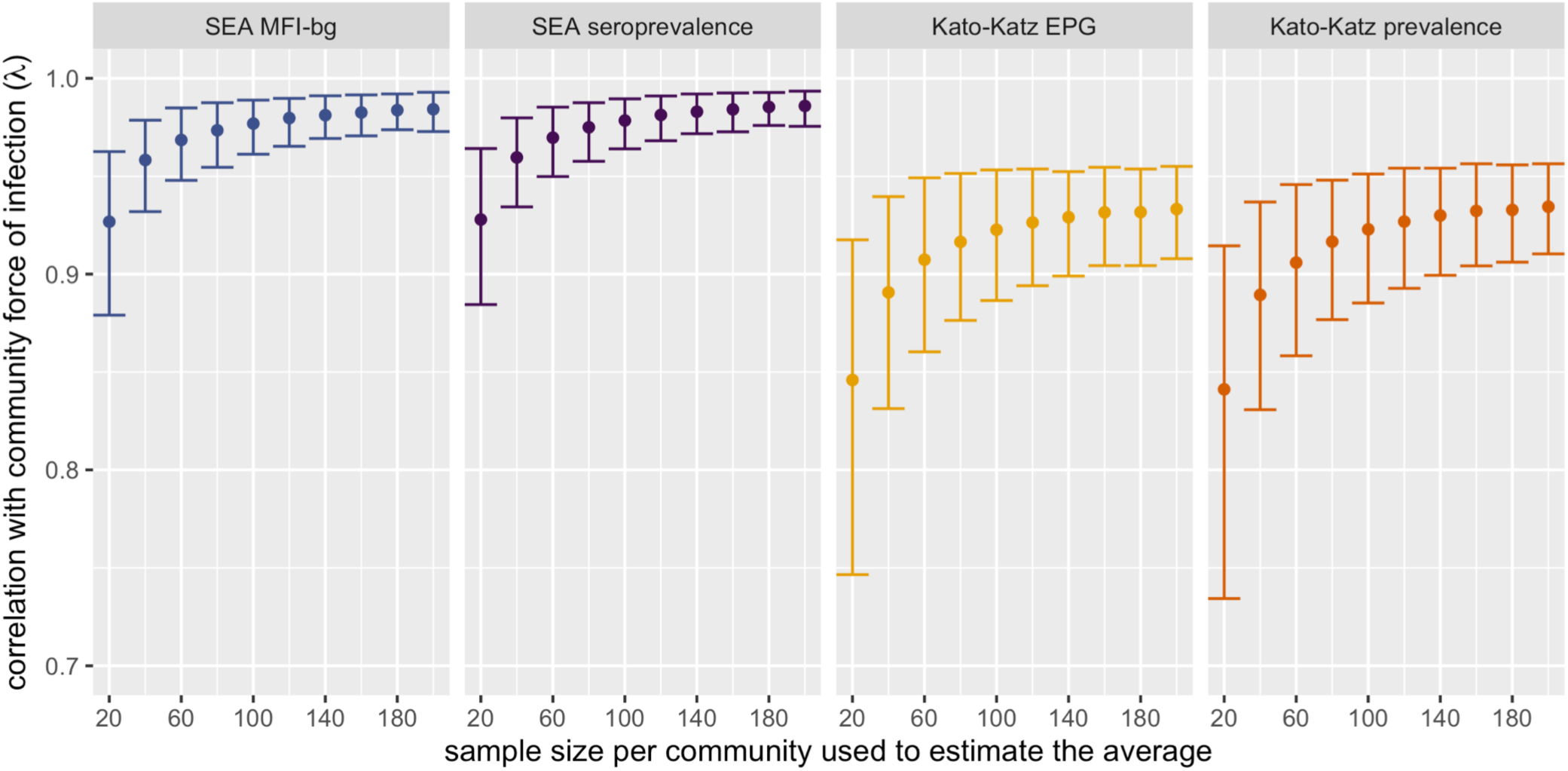
Spearman rank correlaton between measures of *Schisotosoma mansoni* and community level serological force of infection (!) estimated with different simulated sample sizes per community. Force of infection was measured by the seroconversion rate in the full sample (median 117 measurements per community). Estimates and error bars mark bootstrapped means and 95% confidence intervals. In each replicate, samples of different sizes were drawn with replacement from each of 30 villages before calculating the means and correlation with force of infection estimated in the full sample. MFI-bg: median fluorescence intensity minus background on the Luminex platform; SEA: soluable egg antigen; EPG: eggs per gram of stool. Created with notebook: https://osf.io/mhdjx/

### Gradient of transmission with distance from Lake Victoria

Based on studies using stool exam only, we hypothesized that serological measures of transmission intensity would be higher among children living closer to Lake Victoria compared with children living further away from the lake.^21,23^ *S. mansoni* is transmitted to humans through dermal exposure to cercariae in surface water and the predominant exposure in this setting was Lake Victoria. We summarized community-level seroprevalence and force of infection by distance from the lake and found a clear gradient of transmission (Fig 4a, 4b). Among study children, 94% of infections by Kato-Katz and 93% of SEA seropositive preschool age children lived within 1.5 km of Lake Victoria, consistent with highly focal transmission.

**Figure 4.**
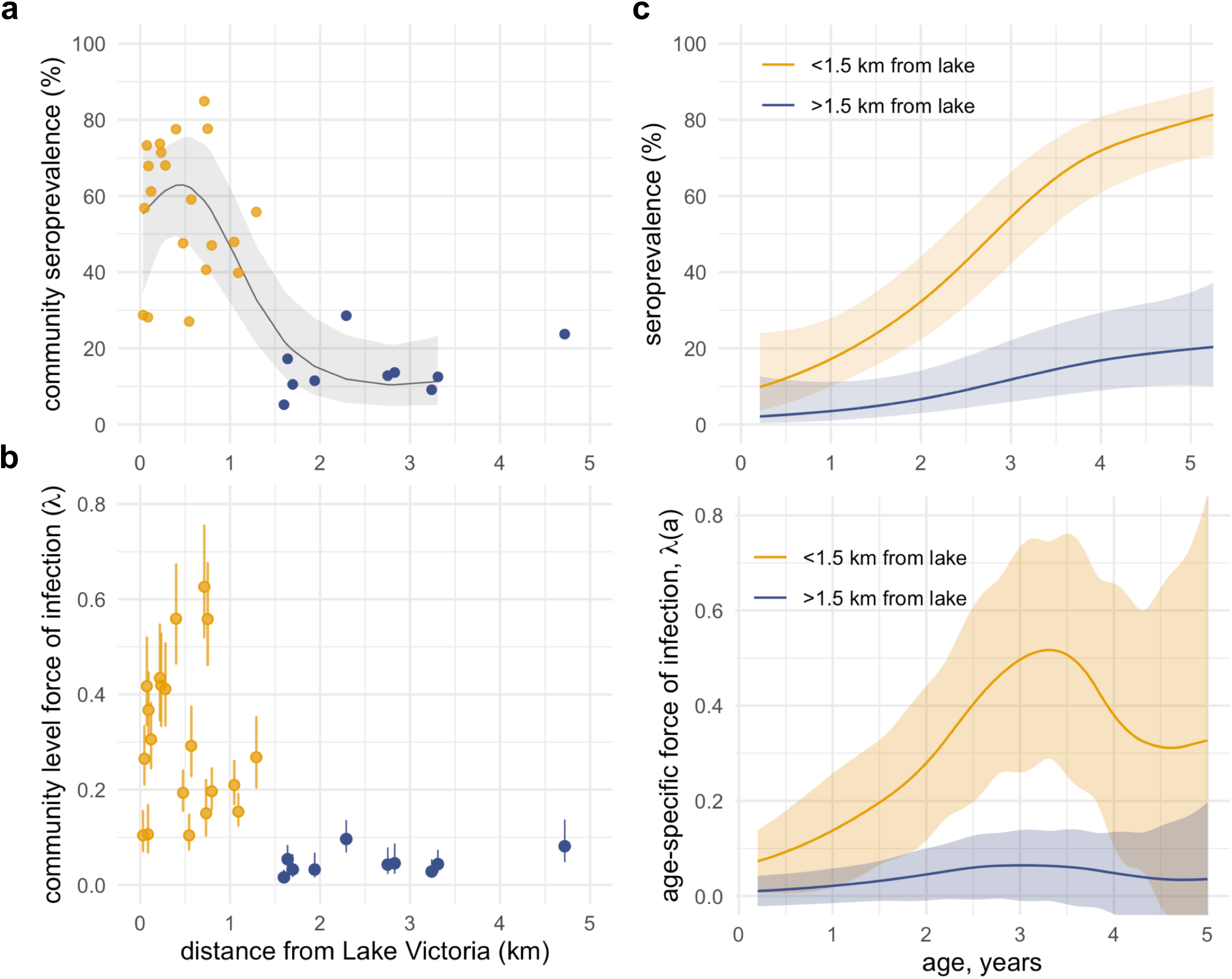
*Schisotosoma mansoni* SEA seroprevalence and force of infection among preschool aged children by distance to Lake Victoria and age in Mbita, Kenya, 2012–2014. (**a**) Relationship between distance from Lake Victoria and village-level SEA seroprevalence in the 30 study communities over the three year period. Cubic spline fit and approximate simultaneous 95% confidence interval summarizes the relationship between distance from the lake and seroprevalence, with predictions up to 3.5 km to reduce edge effects. (**b**) Relationship between distance from Lake Victoria and village-level force of infection estimated by the SEA seroconversion rate in the 30 study communities. Vertical bars on community-level force of infection estimates mark 95% confidence intervals estimated from a generalized linear model. In panels **a** and **b**, points are colored by communities further than 1.5 km from the lake, used to examine differences in age-varying force of infection. (**c**) Age-dependent seroprevalence and force of infection estimated with cubic splines, stratified by distance to Lake Victoria. Shaded regions indicate approximate simultaneous 95% confidence intervals. Created with notebooks: https://osf.io/w7rkz/, https://osf.io/uj75h/.

### Age-varying force of infection

We hypothesized that age-varying force of infection would be higher among children who lived closer to Lake Victoria compared with children who lived further away. We also hypothesized that higher transmission would be reflected in a steeper age-seroprevalence curve that plateaued at a higher level, consistent with seroepidemiologic patterns observed across diverse pathogens ^5,24^ but which has not been previously examined for *S. mansoni*. We stratified villages by those that were within 1.5 km of Lake Victoria versus those further away based on the observed distribution of community-level force of infection, noting that all villages with high seroprevalence and force of infection were <1.5 km from Lake Victoria (not a pre-specified cutoff). We estimated age-varying seroprevalence and force of infection using semiparametric cubic splines that controlled for child age between groups.

Higher transmission intensity among villages close to Lake Victoria was reflected in a steeper age-seroprevalence curve (Fig 4c). By 5 years of age, 80% of children living in villages <1.5 km from Lake Victoria had serological evidence of infection, compared with 20% of children living further from the lake. Age-varying force of infection estimated from age-structured seroprevalence with semiparametric cubic splines showed that force of infection increased through the third year of age (Fig 4c). At age three years, the seroconversion rate among children living < 1.5 km from Lake Victoria was 0.5 per child-year, which means that among 3-year old children who were not previously infected, 50% were infected before age 4 years. A sensitivity analysis by quintiles of distance from the lake showed that there were only two distinct age-seroprevalence profiles that were delineated by the 1.5 km threshold (Supplementary Fig 3).

## Discussion

To our knowledge, this study represents the first comprehensive examination of *S. mansoni* seroepidemiology among children living in endemic settings. Close alignment between measures of transmission intensity derived from stool infections and antibody response provides robust evidence to support the use of well-characterized schistosome antigens to monitor transmission and burden of schistosomiasis in similar settings. In this endemic population, antibody response to *S. mansoni* SEA provided information about transmission at fine spatial scales and at high resolution across ages. All high transmission villages fell within 1.5 km of Lake Victoria, and in this study 94% of infections among preschool age children fell within this narrow band around the lake. Our estimates of age-specific force of infection suggest that infection pressure begins very early and that force of primary infection peaks by age 3–4 years among children living in villages close to the lake.

Our study supports a growing consensus for the need to expand control programs to treat and prevent infection among preschool age children in endemic settings.^25^ In communities along the shores of Lake Victoria, antibody response to SEA showed that 55% of children were infected by age three years, at least two years before they would be eligible to receive treatment through school-based MDA. Global control programs currently exclude preschool age children due in part to lack of evidence for efficacy and safety in this group, but clinical trials are in progress to develop safe delivery methods for praziquantel for children under 6 years old. ^26,27^ Complementary strategies that focus on environmental interventions should be promoted in tandem with MDA to help reduce transmission among this vulnerable age group.^28^

Serological monitoring among older, school age children could be less useful in high transmission settings because SEA IgG remains high in infected individuals and decays very slowly after treatment, even in the absence of reinfection. In this study, >80% of children were seropositive by age five years in villages close to the lake. If SEA IgG levels remain elevated for multiple years following infection, then a majority of school-age children will be seropositive and changes in transmission would only be detectable amongst children born into lower transmission conditions. Longitudinal measurement of antibody levels to SEA among children born in the same setting in western Kenya as the present study showed maternal IgG clearance by age 20 weeks and no evidence for antibody waning among a limited number of children infected through age 24 months.^29^ Longer, longitudinal follow-up of children who have been successfully treated and not re-infected would help confirm SEA IgG decay rates, but clearly delineated, wide separation between SEA antibody responses at low and high levels observed in this study (Supplementary Fig 4) provides indirect evidence for a robust and durable IgG response with little waning among preschool age children.

A shift in age-seroprevalence curves for *S. mansoni* with reductions in transmission closely aligns with patterns observed for many pathogens,^24^ and aligns with the “peak shift” of infection intensity observed for schistosomiasis and many other parasitic diseases.^30,31^ That *S. mansoni* SEA exhibits this classic seroepidemiologic pattern supports the use of modeling approaches originally developed in the context of vaccine preventable diseases and arboviruses to study transmission through cross-sectional serological surveys.^32^ The use of widely generalizable modeling approaches to estimate *S. mansoni* transmission in cross-sectional, serological surveys further support its inclusion in integrated surveillance platforms, which benefit from shared analysis methods to translate antibody response into measures of transmission.^18^ Multiplex antibody testing from a lymphatic filariasis surveillance survey in coastal Kenya recently identified villages with a clear signature of *S. mansoni* transmission based on SEA seroprevalence,^33^ The methods we have used herein, such as model-based geostatistical mapping and force of infection estimation, could further extend analysis outputs of integrated serosurveillance platforms to help inform control programs.

In addition to broader surveillance, serology could directly inform the design and evaluation of schistosomiasis intervention studies. For example, some villages near Lake Victoria have higher forces of transmission and are less responsive to MDA than others. Heterogeneity in transmission and responsiveness to intervention emerges as early as 2 years after control program initiation,^34,35^ but our results suggest that a single cross-sectional serological survey before programs start could estimate community-level seroprevalence or force of infection and provide guidance for which villages warrant more intensive intervention. Furthermore, antibody responses among preschool age children may be useful to reflect the impact of an intervention, thereby improving monitoring and evaluation.

This analysis had limitations. The 30 study villages were located over a relatively small area (277 km^2^). We were unable to test whether spatial predictions of *S. mansoni* seroprevalence remain accurate if sampling locations are more widely distributed over larger areas, such as districts or nations. Fine-scale heterogeneity in force of infection observed in this study suggests that program monitoring at the level of districts might be too coarse as populations approach elimination, but surveys with high spatial resolution over larger geographic areas could provide confirmatory results. Strong relationships between remotely sensed covariates and measures of *S. mansoni* infection observed in this study (Supplementary Fig 5) suggest it might be possible to use geospatial models to predict seroprevalence over larger areas with less densely measured communities, but it remains an open question. It might also be possible to improve geostatistical model predictions by extending models to include more geospatial layers ^21^ or by including more refined measures of snail habitat derived from remotely sensed images, such as those recently proposed in Senegal.^36^ The multiplex antibody assay used to measure SEA response in this study is not commercially available and there is currently no rapid diagnostic test based on SEA. At this time, large-scale monitoring of SEA response is therefore most viable through integrated serologic surveillance with specimens tested in multiplex in larger reference labs, such as the Kenya Medical Research Institute (KEMRI) laboratory in Nairobi where the present samples were analyzed. The force of infection analyses assumed no seroreversion over the age range, and longitudinal studies of children following infection and successful treatment without reinfection would be useful to confirm the post-infection SEA IgG kinetics. Finally, since SEA IgG appears to be a durable response, it cannot detect reinfection or superinfections; thus, force of infection estimates based on SEA provide a lower bound of the true transmission intensity.

In conclusion, antibody response to *S. mansoni* SEA antigen among preschool age children enabled fine-scale estimates of transmission heterogeneity in an endemic setting in western Kenya. Serological estimates aligned with those from stool-based testing and provided higher resolution about between-village heterogeneity in infection than microscopy at lower levels of transmission. Our results support the inclusion of *S. mansoni* in population-based serologic surveillance platforms to target control programs and monitor their effectiveness.

## Materials and Methods

### Ethics Statement

The study protocol was reviewed and approved by ethical review committees at the Kenya Medical Research Institute (KEMRI, SSC number 2185) and the U.S. Centers for Disease Control and Prevention (protocol 6249). The analysis protocol was further reviewed and approved by the ethical review committee at the University of California, San Francisco (protocol 19-28772). The study was explained to participants and field staff obtained written, informed consent at the time of enrollment in each study year. Parents or guardians provided consent for the preschool age children enrolled.

### Study design

Thirty villages were enrolled in a community-based, randomized, and controlled trial that delivered annual praziquantel MDA to primary school children (school-based delivery, 15 communities) or community-wide (all people 5 years or older, 15 communities). Villages that had *S. mansoni* prevalence of ≥25% prevalence by Kato-Katz in an earlier survey of the study region^20^ were selected to participate in the trial. At the time of the study, there were no guidelines for the inclusion of preschool age children in schistosomiasis MDA programs, so only preschool age children identified as positive for *S. mansoni* infection by Kato-Katz were treated with crushed praziquantel under medical supervision. Additional details of the trial and study population have been previously reported.^19^ All children ages 1 to 5 years old were eligible for study inclusion and were enrolled in separate cross-sectional surveys in May to July of 2012, 2013, and 2014. A small number of children between ages 2 months and 1 year (*n* = 51) and slightly older than 5 years (*n* = 158) were enrolled outside of the target age range and were included in the present analysis.

### Specimen collection and testing

Eligible children provided stool and blood at a central location in each community. Plastic stool containers were given to the parents and an attempt was made to collect a single stool sample from the child at each village’s central monitoring location. Stool samples were transported in cool boxes with ice packs to the Ministry of Health’s Division of Vector-Borne Diseases (DVBD) laboratory in Homabay, where they were processed the same day. From each stool sample, two slides were prepared by the Kato-Katz method and examined by independent, trained microscopists for the presence of *S. mansoni* eggs. Arithmetic means of the two slides were calculated and expressed as eggs per gram (EPG). The average EPG values from the two readers were used in the analysis.

Blood from a fingerstick was collected into a serum capillary collection tube (approximately 100 μL) and transported to the DVBD laboratory in Homabay for centrifugation. Sera were stored at −20°C in the DVBD laboratory and were transported to the KEMRI NTD laboratory in Kisumu, where they were stored at −80°C until sent to KEMRI/ESACIPAC laboratory in Nairobi. Specimens were analyzed on the Luminex platform (Bio-Plex 200) for antibody levels to the *S. mansoni* soluble egg antigen (SEA) and recombinant Sm25 antigen, an integral glycoprotein found in microsomal preparations of *S. mansoni* adult worms (GenBank Accession M37004.1).^19^ Details of the assay preparation have been previously reported.^19^ Samples were tested in duplicate and those with a coefficient of variation of >15% between duplicate wells for >3 positive antibody responses were repeated.

In this analysis, we focused on SEA antibody responses because Sm25 responses did not clearly distinguish two subpopulations of seronegative and seropositive (infected or previously infected) children (Supplementary Fig 4). A SEA seropositivity cutoff was determined through a receiver operating characteristic curve analysis of known positive and negative sera with a cutoff for the KEMRI/ESACIPAC instrument determined as 965 MFI-bg (sensitivity = 97.5%, specificity = 100%) as previously described.^19^

### Prevalence mapped with model-based geostatistics

For each location in the study region, we calculated the distance from Lake Victoria using the global surface water layer,^37^ and obtained average monthly minimum temperatures and average monthly precipitation from WorldClim.^38^ We considered including elevation in the geostatistical model but omitted it because it was highly correlated with distance from the lake (Supplementary Fig 5). We modeled prevalence as a function of distance to the lake, temperature, and precipitation using a spatial generalized additive model with binomial error structure and cubic splines for each covariate. The model included a spatially structured, village-level random effect with a low rank Gaussian Process and Matern covariance structure, implemented in the R mgcv package.^39^ We made predictions at 1 km resolution (minimum scale of the WorldClim data) and summarized observed versus predicted prevalence at the 30 village locations.

### Community level measures of infection

We estimated community level prevalence of *S. mansoni* infection as the proportion of children with detected eggs in stool. We estimated community mean eggs per gram on the log10 scale because infection intensities were highly skewed. For children without any eggs detected in stool we recoded them as 1 to enable the log10 transformation before estimating community means. Thus, mean eggs per gram is reported as the overall community average infection intensity, not the average infection intensity among infected children. We compared community level estimates of seroprevalence, force of infection (seroconversion rate, described below), Kato-Katz prevalence, and mean eggs per gram of stool using bivariate scatter plots with locally weighted regression smooths and Spearman rank correlation. We estimated the Spearman rank correlation of community level seroprevalence across study years to assess the stability of estimates over time. We did not adjust community level seroprevalence or infection prevalence for age because the age distribution was very similar across all villages (Supplementary Fig 6).

We conducted a simulation study to assess the effect of community-level sample size on the relationship between multiple SEA and Kato-Katz measures and community-level force of infection measured by the SEA seroconversion rate. The objective of the simulation was to determine whether a small number of measurements in a community could still provide rank-order information about community-level force of infection. From the empirical measurements, we resampled children with replacement from each village using a non-parametric bootstrap with village-level sample sizes that varied from 20 to 200 measurements per village. In each bootstrap replicate, we estimated the mean log10 SEA MFI-bg, SEA seroprevalence, mean log10 Kato-Katz eggs per gram, and Kato-Katz prevalence, and then estimated the Spearman rank correlation between each measure and community-level force of infection measured by the seroconversion rate in the full sample. We used the 2.5% and 97.5% percentiles of the bootstrap distribution to estimate 95% confidence intervals for correlation estimates at each sample size.

### Community level force of infection by distance from Lake Victoria

We estimated community level force of infection from age structured seroprevalence assuming a constant force of infection over different ages, *λ*(*a*) = *λ* and no seroreversion.^40^ We modeled probability of being seropositive as a function of child age with a generalized linear model fit within each community that assumed a binomial error structure and complementary log-log link:

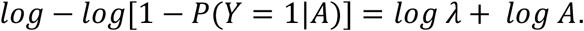

We estimated the community level force of infection and its 95% confidence interval from the model’s maximum likelihood estimate of the intercept term.^41^

### Age Structured seroprevalence and force of infection

We extended the community-level force of infection model to allow force of infection to vary by age using cubic splines in a generalized additive model.^42^ We fit a generalized additive mixed model with binomial error structure and logit link:

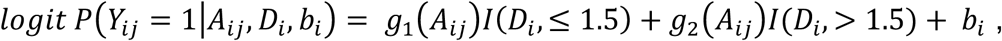

where *Y*_*ij*_ is SEA seropositivity and *A*_*ij*_ is age for child *j* in village *i. D*_*i*_ is distance from Lake Victoria to community *i*. Community-level random effects, *b*_*i*_, were included to allow for correlated outcomes within community. Functions *g*_1_(·), *g*_2_(·) were parameterized with cubic splines that had smoothing parameters chosen through cross-validation and were allowed to differ between villages <1.5 km from Lake Victoria versus further away.

We estimated age-specific seroprevalence from the model as:

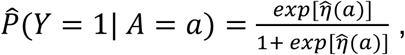

and we estimated age-specific force of infection from the model as:

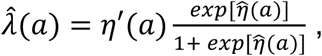

where *η*’(*a*) is the first derivative of the linear predictor from the logit model, *η*(*a*).^43^ We estimated *η*’(*a*) using finite differences from the generalized additive mixed model predictions.^13,39,42^ We estimated approximate, simultaneous 95% confidence intervals around age-specific seroprevalence and age-specific force of infection curves with a parametric bootstrap (10,000 replicates) from posterior estimates of the model parameter covariance matrix.^44^

## Data Availability

Data and replication files required to conduct the analyses are available through GitHub and the Open Science Framework: https://osf.io/rnme8. Community location data required to replicate the geostatistical model fits is not publicly available to protect participant confidentiality but is available from the corresponding author upon request, pending appropriate human subjects review and approval.

## Acknowledgements

This work was supported by the National Institutes of Allergy and Infectious Diseases (K01 AI119180 to BFA) and the Bill & Melinda Gates Foundation (OPP1022543 to PJL). The findings and conclusions in this article are those of the authors and do not necessarily represent the official position of the Centers for Disease Control and Prevention. Use of trade names is for identification only and does not imply endorsement by the Public Health Service or by the U.S. Department of Health and Human Services.

## Author contributions

**CRediT Taxonomy:**

**Conceptualization:** BFA, PJL, KYW, MRO

**Data Curation:** KYW, MRO, BFA

**Formal Analysis:** BFA

**Funding Acquisition:** BFA, PJL

**Investigation:** MRO, KYW, HK, SMN, FOR, JWP, WES

**Methodology:** BFA

**Project Administration:** MRO, KYW

**Resources:** HK, SMN, JWP

**Software:** BFA

**Supervision:** MRO, SMN, PJL, WES

**Validation:** JWP, SMN

**Visualization:** BFA

**Writing – Original Draft Preparation:** BFA, MRO

**Writing – Review & Edition:** BFA, HK, SMN, FOR, JWP, WES, PJL, KYW, MRO

---

**Supplementary Figure 1.**
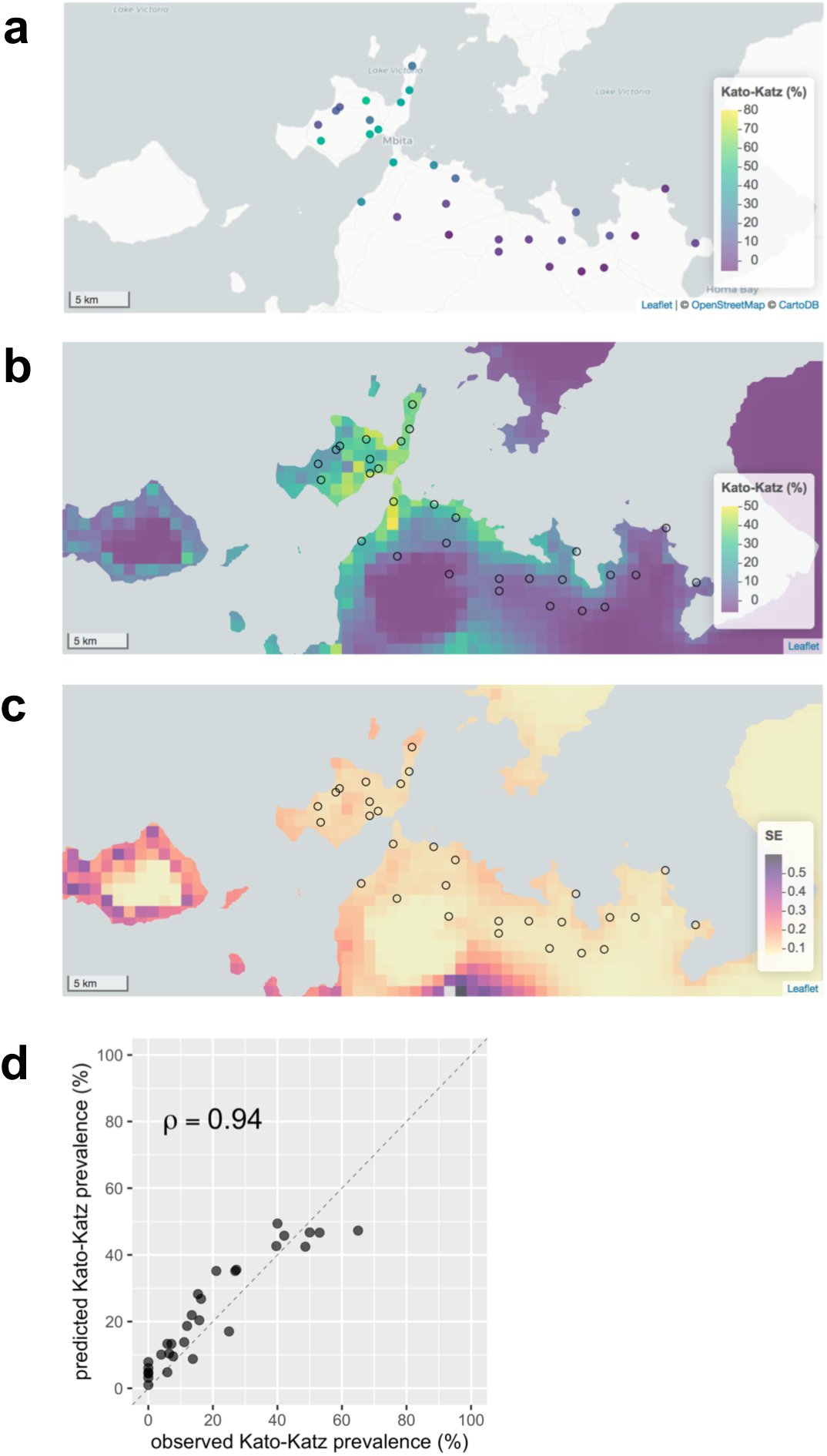
Spatial heterogeneity of *Schisotosoma mansoni* infection prevalence in Mbita, Kenya, 2012–2014. Infection measured using dual slide Kato-Katz microscopy in 3,426 stool specimens among pre-school aged children. (**a**) Infection prevalence in the 30 study communities. (**b**) Predicted prevalence at 1 km resolution from a geostatistical model. (**c**) Approximate standard errors of the predicted proportion infected from the geostatistical model. (**d**) Predicted versus observed infection prevalence for the 30 study villages in 2014 and spearman rank correlation (*ρ*). The diagonal line is 1:1. Created with notebook: https://osf.io/eqp3w/.

**Supplementary Figure 2.**
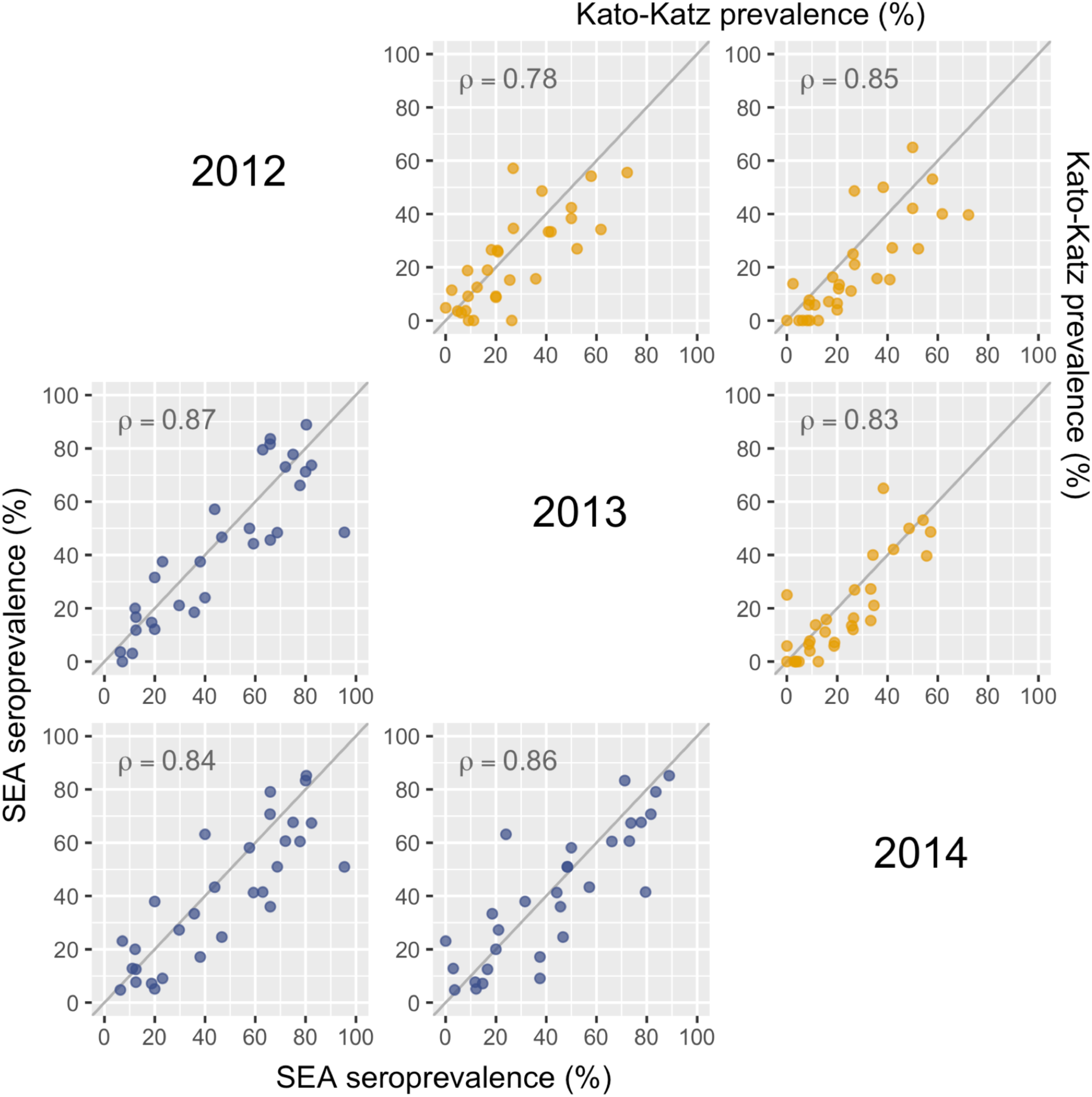
Relationship between community level SEA seroprevalence and Kato-Katz infection prevalence across years. Bivariate pairs plot of SEA seroprevalence (lower panels) and Kato-Katz infection prevalence (upper panels) by year in the 30 study communities, including Spearman rank correlation estimates (*ρ*). Each panel shows the relationship between community level prevalence between years on the diagonal. Created with notebook: https://osf.io/mhdjx/.

**Supplementary Figure 3.**
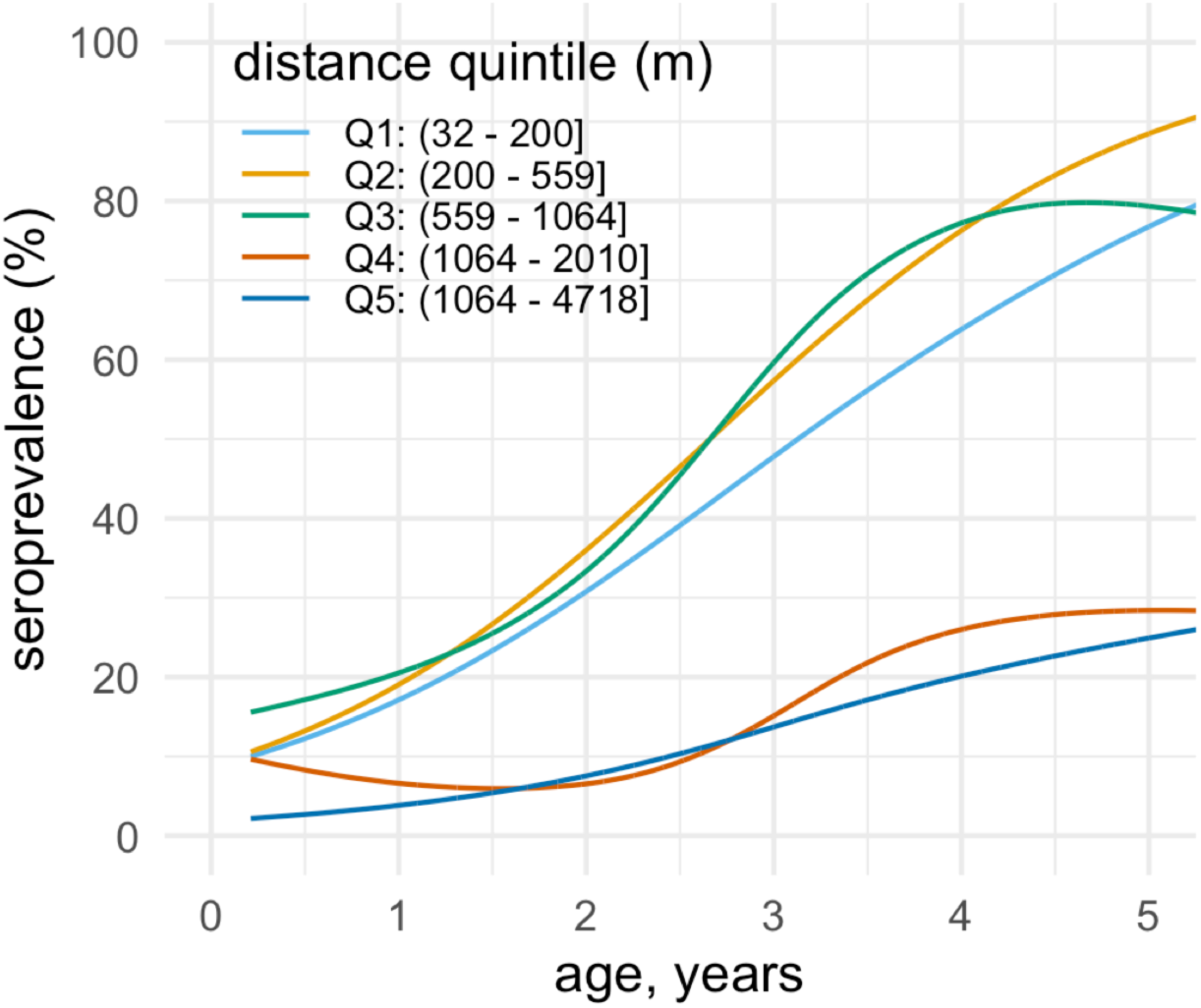
Seroprevalence of *Schisotosoma mansoni* SEA by age and qunitile of distance from Lake Victoria, Mbita, Kenya, 2012–2014. Age dependent seroprevalence was estimated using semiparametric cubic splines as in the primary analysis. Each distance quintile includes measurements from six communities (*n*=3,663 measurements from 30 villages). Created with notebook: https://osf.io/uj75h/.

**Supplementary Figure 4.**
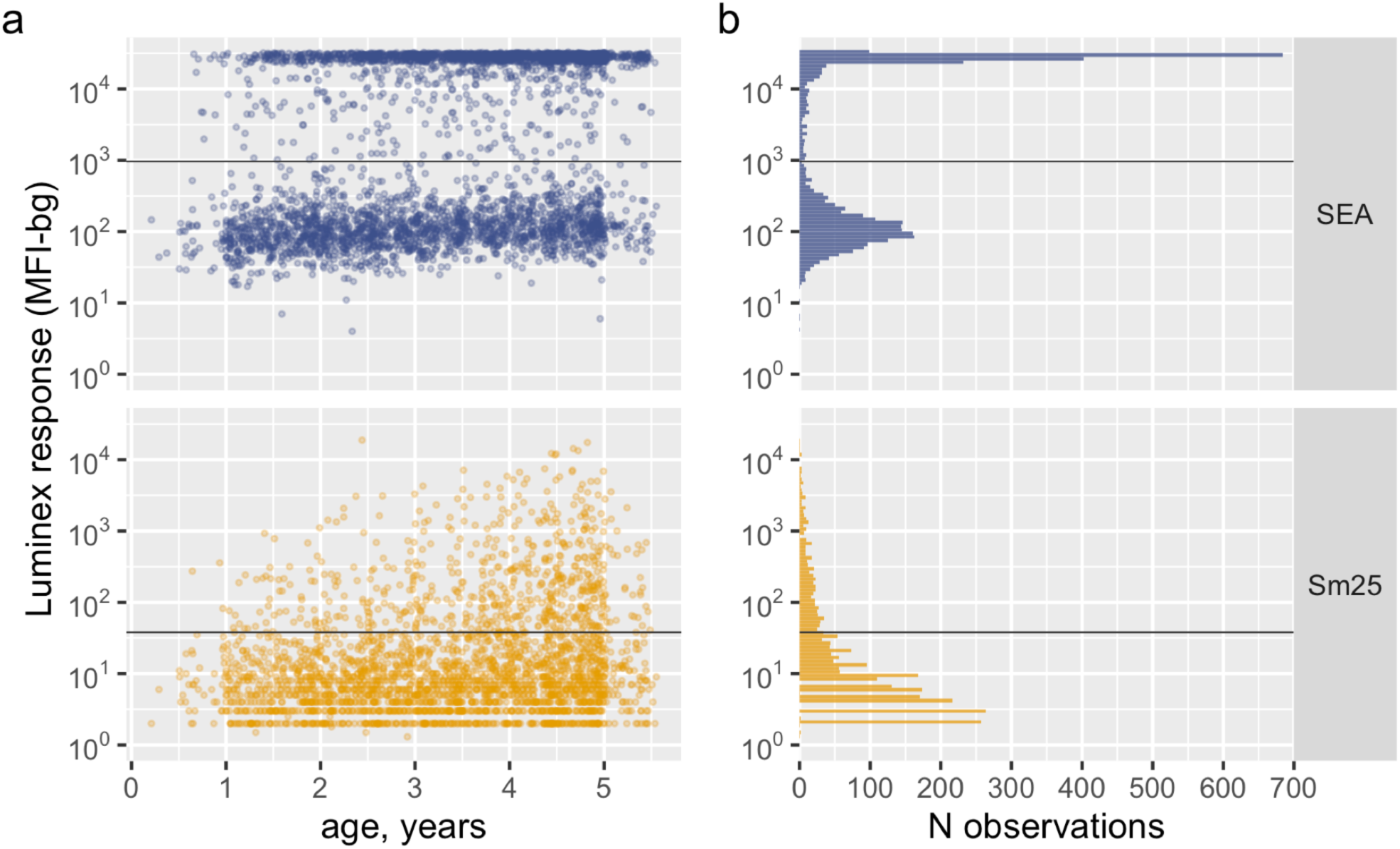
Antibody responses to *Schisotosoma mansoni* SEA and Sm25 antigens from 3,663 preschool aged children near Lake Victoria, Mbita, Kenya, 2012–2014. Luminex response in by age. (**b**) Distribution of Luminex responses. In both panels, the horizontal line marks the seropositivity cutoff derived through receiver operating characteristic curve analyses of known positive and negative samples (SEA: 965 MFI-bg, sensitivity = 97.5%, specificity = 100%; Sm25: 38 MFI-bg, sensitivity = 93.5%, specificity = 97.3%). MFI-bg: median fluorescence intensity minus background. Created with notebook: https://osf.io/gdhb6/.

**Supplementary Figure 5.**
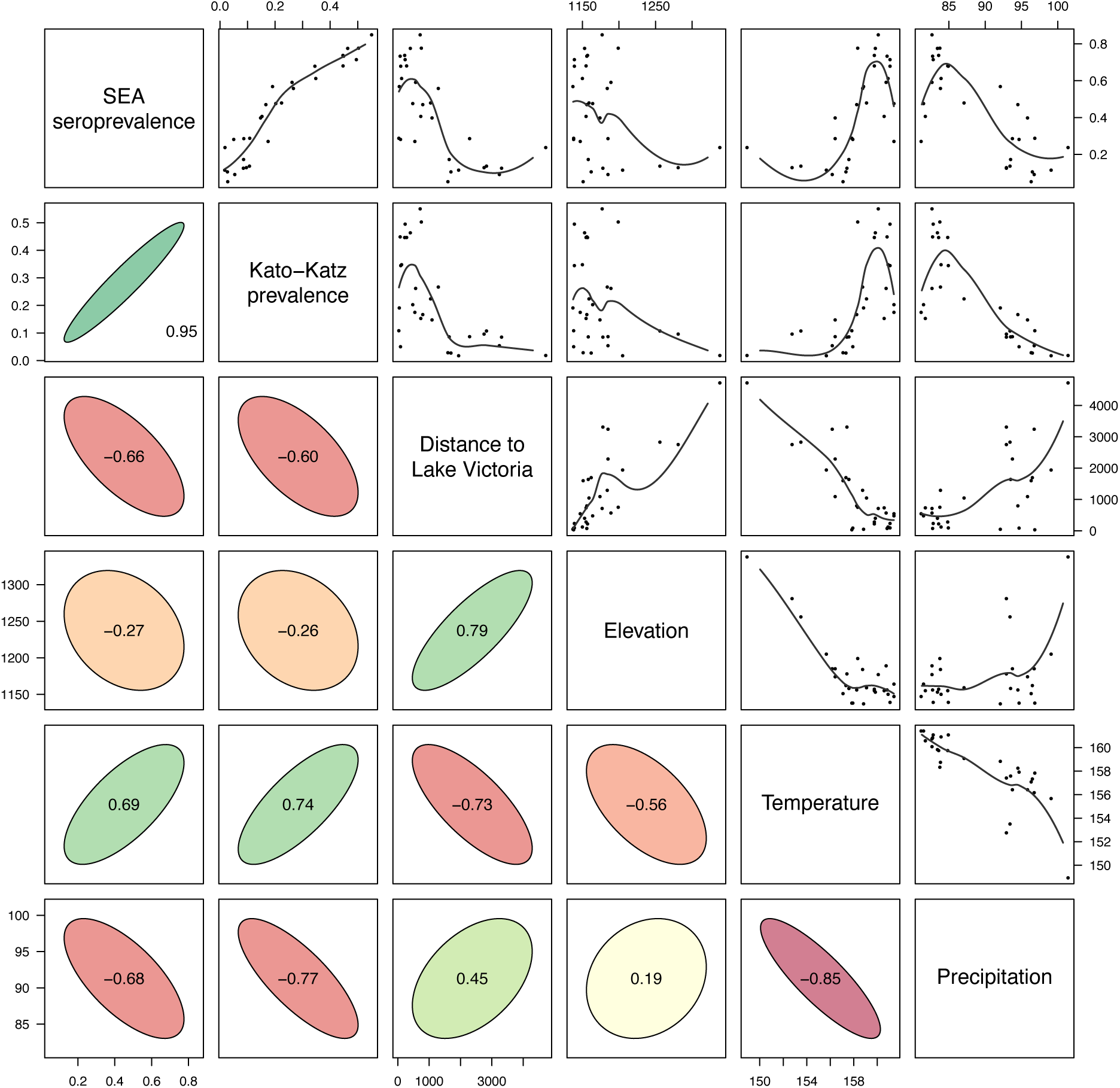
Bivariate pairs plot of *Schisotosoma mansoni* SEA seroprevalence, Kato–Katz prevalence, and environmental covariates in 30 communities near Lake Victoria, Mbita, Kenya, 2012–2014. Scatter plots include nonparametric locally weighted regression fits trimmed to reduce edge effects. Correlation ellipses depict the strength of the association on the basis of the Spearman rank correlation (printed). Preschool aged children < 5 years old were measured for SEA seroprevalence (*N* = 3,663) and Kato–Katz prevalence (*N* = 3,426). Created with notebook: https://osf.io/eqp3w/.

**Supplementary Figure 6.**
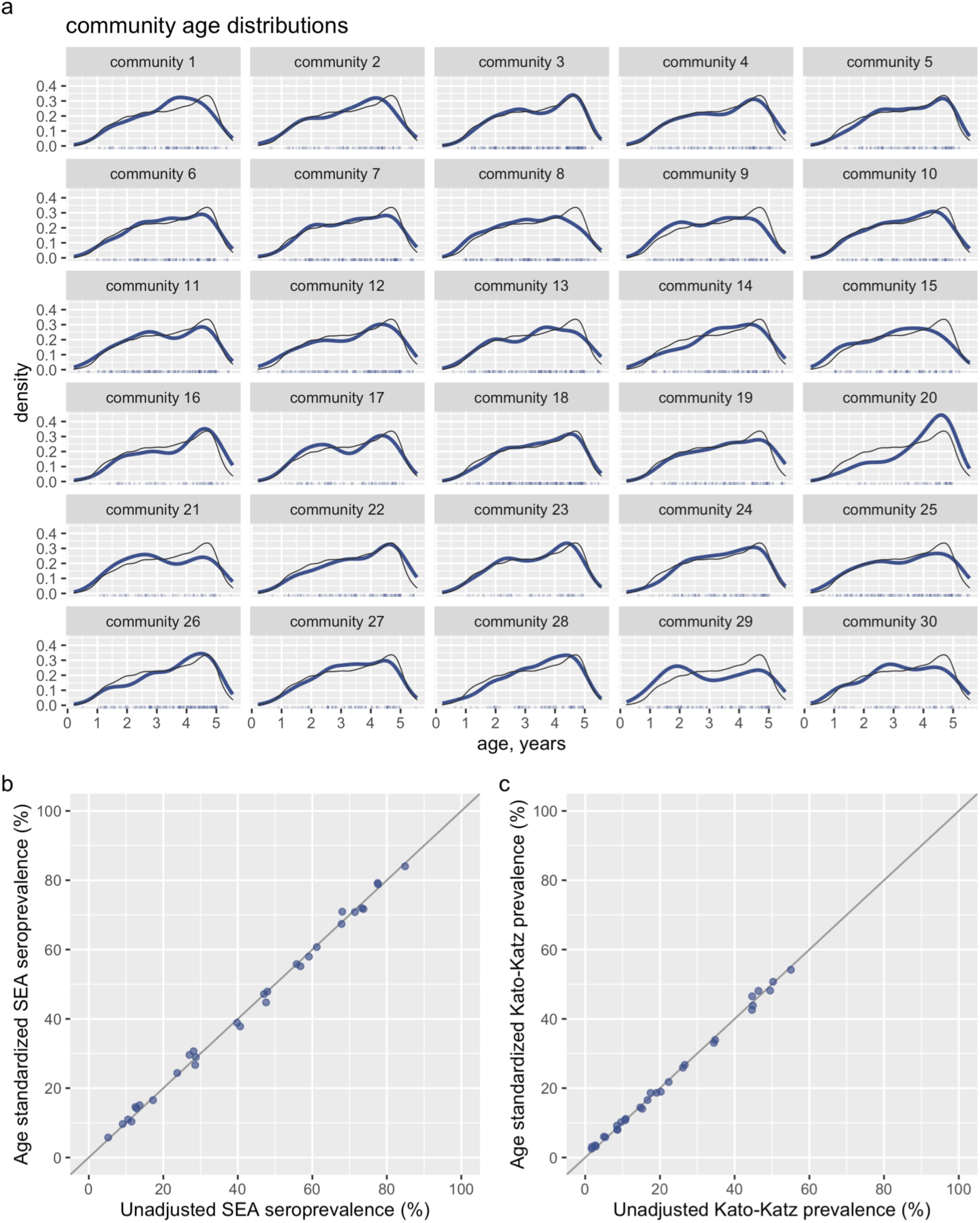
Age distribution and effect of age adjustment on community level prevalence estimates based on 3,663 samples collected in 30 communities near Lake Victoria, Mbita, Kenya, 2012–2014. (**a**) Age distribution in each community. The heavy line is each community’s age distribution, the light grey line marks the age distribution across all communities. Age standardized community level seroprevalence to *Schistosoma mansoni* SEA antigen versus unadjusted seroprevalence; the diagonal line is 1:1. (**c**) Age standardized community level infection prevalence to *S. mansoni* measured with dual slide Kato-Katz microscopy versus unadjusted prevalence; the diagonal line is 1:1. Created with notebook: https://osf.io/w7rkz/.

